# Relationship among work–treatment balance, job stress, and work engagement in Japan: A cross-sectional study

**DOI:** 10.1101/2021.10.23.21265407

**Authors:** Kazunori Ikegami, Hajime Ando, Hisashi Eguchi, Mayumi Tsuji, Seiichiro Tateishi, Koji Mori, Keiji Muramatsu, Yoshihisa Fujino, Akira Ogami, for the CORoNaWork Project

## Abstract

There is a drive to support workers undergoing medical treatment who wish to continue working in Japan, known as the work–treatment balance. It is hoped that this support for the work–treatment balance could boost their mental health. This study examines the relationship among the work–treatment balance, job stress, and work engagement. This study was conducted in December 2020 in Japan, with 27,036 participants. We divided the participants into three groups by the receipt state of support for the work–treatment balance: control group (no need the support), unsupported group, and supported group. The scores of the parameters of the job content questionnaire and the Utrecht Work Engagement Scale (UWES-3) were compared among groups using a multilevel regression with age-sex or multivariate-adjusted models. In the two models, the job control score of the unsupported group was significantly lower than that of the control group. The two social support scores of the supported group were significantly higher than those of the control group. The scores of the UWES-3 of the unsupported group were significantly lower than those of the control group. The support of work–treatment balance for workers could have a positive impact on their mental health.

## INTRODUCTION

Fitness to work (FTW), the process of assuring that an employee can complete a task without risk to their health and safety or those of others, is one of the occupational health issues. In Japan, support for the return to work (RTW) or FTW in the workplace has been promoted for workers with mental health disturbances such as depressive disorders since early 2000. ^1, 2)^ In recent years, systems to support balance between working life and medical treatment (the work–treatment balance) have been promoted to reinforce FTW for workers with various chronic diseases, including cancer, brain disease, and intractable diseases. There is a widespread movement to support workers willing to continue working while receiving treatment for their chronic diseases. ^3)^

Support for work–treatment balance is promoted through cooperation among workers, workplace staff, occupational physicians, and the attending physician. When sharing information among these stakeholders, employment considerations such as changing the work location or the work content and shortening the working hours according to the worker’s condition will be considered. In addition, the support related to the mental health of workers receiving support for work–treatment balance is considered important. ^3)^

The coronavirus disease 2019 (COVID-19) pandemic has had a major impact on health, life style, and work. ^4-6)^ People with chronic diseases are concerned about the risk of severe acute respiratory disease due to the COVID-19 infection. ^7, 8)^ Additionally, they have faced the risk of their diseases worsening due to interruption of treatment, or have suffered a deterioration of their physical and mental health status due to numerous restrictions on daily living and work practice related to COVID-19, including social distancing and self-quarantine. ^9, 10)^ From these aspects, the work–treatment balance of workers with chronic diseases could be important for reducing mental distress by enabling them to continue work.

In this study, we focused on the work–treatment balance and job stress and hypothesized that workers who can receive support for the work–treatment balance will have lower job stress and higher work engagement. We used data from the Collaborative Online Research on Novel-coronavirus and Work study (CORONaWork study) to clarify the relationship among the work–treatment balance, job stress, and work engagement.

## SUBJECTS AND METHODS

### Study design and setting

We conducted a prospective cohort study by a research group consisting of the University of Occupational and Environmental Health, the CORoNaWork study. This study was conducted as a self-administered questionnaire survey by a Japanese Internet survey company (Cross Marketing Inc. Tokyo) from December 22 to 25, 2020. Incidentally, during the baseline survey, the number of COVID-19 infections and deaths were overwhelmingly higher than in the first and second waves; therefore, Japan was on maximum alert during the third wave.

This study design is a cross-sectional study using a part of a baseline survey of the CORoNaWork study. Fujino et al. introduced the details of this study protocol ^11)^.

### Participants

Participants were aged between 20 and 65 and were working at the time of the baseline survey. A total of 33,087 participants, who were stratified by cluster sampling by gender, age, region, and occupation, participated in the CORoNaWork study. Of this total number, only 27,036 responses were eligible for the analysis.

### Questionnaire

The questionnaire items used in this study were described in detail by Fujino et al. ^12)^. We used questionnaire data on sex, age, educational background, area of participants’ residence, occupation, company size where participants work, working hours per day, family structure, the receipt state of the support for the work–treatment balance, work-related questionnaires like the Japanese version of the Job Content Questionnaire (JCQ) ^13, 14)^, and the three-item Japanese version of the Utrecht Work Engagement Scale (UWES-3) ^15, 16)^.

Regarding the receipt state of the support for the work–treatment balance, we asked, “have you received any support from your company to continue working in your current health condition?.” The responses were the following three options: Not necessary (those who do not need that support), No, I do not receive despite I need the support (those who need that support but were not receiving it), and Yes, I do (those who needed that support and were receiving it).

The JCQ, developed by Karasek, is based on the job demands–control (or demand–control–support) model^13)^. The reliability and validity of the Japanese version of the JCQ were demonstrated by Kawakami et al. ^14)^. We used a shortened version of the 22 items in the JCQ, in which each item was rated on a 4-point scale (1 = strongly disagree, 4 = strongly agree). The JCQ includes a five-item job demands scale (score range 12–48, Cronbach’s alpha in the present sample = 0.63), a nine-item job control scale (score range 24–96, Cronbach’s alpha in the present sample = 0.74), a four-item supervisor support scale (score range 4–12, Cronbach’s alpha in the present sample = 0.94), and a four-item coworker support scale (score range, 4–12; Cronbach’s alpha, 0.90).

The three-item Japanese version of the Utrecht Work Engagement Scale (UWES-3) was used to assess work engagement ^15, 16)^. The items of the UWES-3 were selected from among those included in the UWES-9. The UWES-3 has been validated in five countries, including Japan, and includes measures of vigor (one item), dedication (one item), and absorption (one item), with each item measured on a seven-point response scale ranging from 0 (never) to 6 (always/every day). Overall scores on the UWES-3 (range: 0–6) were calculated by averaging the individual item scores.

### Variable

We used the scores of the four parameters of the JCQ and UWES-3 as outcome variables. We divided the participants into three groups according to the receipt state of support for the work–treatment balance: control group (those who do not need support), unsupported group (those who need support but were not receiving it), and supported group (those who needed support and were actually receiving it). These variables were used as the exposure variables.

The following items, surveyed using a questionnaire, were used as confounding factors. Sex, age (20-29yr, 30-39yr, 40-49yr, 50-59yr, ≥60 years), and educational background (junior or senior high school, junior college or vocational school, university, or graduate school) were personal characteristics. Occupation (regular employees, managers, executives, public service workers, temporary workers, freelancers or professionals, others), company size where participants worked (≤9 employees, 10-49, 50–99, 100-499, 500-999, 1,000-9,999, ≥10,000), working hours per day (<8h/d, 8≤ and<9h/d, 9≤and<11h/d, ≥11h/d) were used as work-related factors. In addition, the prefecture of participants’ residence participants was used as another variable.

### Statistical method

To analyze the relationships between the four scales of the JCQ or UWES-3 and the three groups according to the receipt state of the support for the work–treatment balance, we used a multilevel mixed-effects regression with the two models nested in the prefecture of residence as random effects. The two models were analyzed for each predictor variable. In the age-sex adjusted model, we treated the three groups, age, and sex as fixed effects and treated the prefecture of residence as random effects. In the multivariate model, we added educational background as personal characteristics, occupation, company size where participants work, working hours per day as work-related variables to the fixed effects of the age-sex adjusted model. In all tests, the threshold for significance was set at P<0.05. We used Stata/SE Ver.15.1 (StataCorp LLC, Station College, TX, USA) for statistical analyses.

## RESULTS

### Participants and descriptive data

A total of 20,261 participants answered that they did not need any support for the work–treatment balance because of their current good health condition. A total of 4,298 answered that they needed support for the work–treatment balance but were not receiving it, and 2,477 answered that they needed the support and were receiving it (Fig. 1).

**Figure 1.**
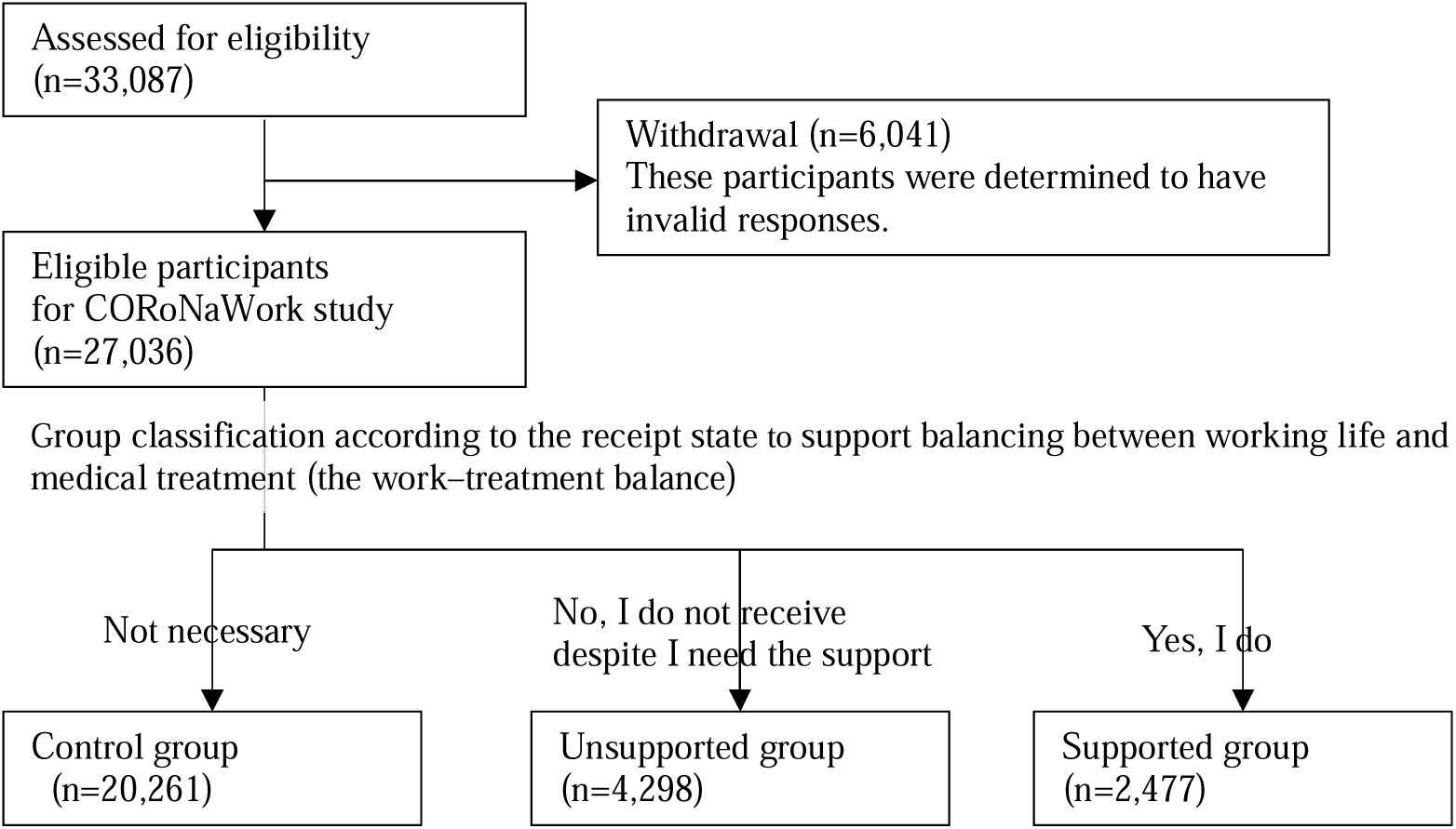
Flow chart of this study population selection.

Table 1 displays the characteristics of the three groups according to the receipt state of support for the work–treatment balance. The supported group had a high proportion of women, college graduates, and those working less than 9 hours a day, and those belonging to company size with ≥1,000 employees. The unsupported group was the proportion of junior high school or high school graduates and those belonging to a company size between 10 and 500 employees.

**Table 1.** Participants’ characteristics in each group according to the receipt state of the support for the work–treatment balance.

| Items | Total |  | Groups by the receipt state of<br>the support for the work–treatment balance |  |  |  |  |  |
| --- | --- | --- | --- | --- | --- | --- | --- | --- |
|  |  |  | Control |  | Unsupported |  | Supported |  |
|  | n (%) / M(SD) |  | n (%) /M (SD) |  | n (%) /M (SD) |  | n (%) /M (SD) |  |
| n | 27036 | (100.0) | 20261 | (100.0) | 4298 | (100.0) | 2477 | (100.0) |
| Sex |  |  |  |  |  |  |  |  |
| Male | 13814 | (51.1) | 10476 | (51.7) | 2171 | (50.5) | 1167 | (47.1) |
| Female | 13222 | (48.9) | 9785 | (48.3) | 2127 | (49.5) | 1310 | (52.9) |
| Generation |  |  |  |  |  |  |  |  |
| 20-29yr | 1905 | (7.0) | 1352 | (6.7) | 316 | (7.4) | 237 | (9.6) |
| 30-39yr | 4858 | (18.0) | 3481 | (17.2) | 817 | (19.0) | 560 | (22.6) |
| 40-49yr | 8011 | (29.6) | 5980 | (29.5) | 1334 | (31.0) | 697 | (28.1) |
| 50-59yr | 9012 | (33.3) | 6809 | (33.6) | 1457 | (33.9) | 746 | (30.1) |
| ≥60yr | 3250 | (12.0) | 2639 | (13.0) | 374 | (8.7) | 237 | (9.6) |
| Educational background |  |  |  |  |  |  |  |  |
| Junior or senior high schools | 7321 | (27.1) | 5477 | (27.0) | 1218 | (28.3) | 626 | (25.3) |
| Junior college or vocational school | 6544 | (24.2) | 4826 | (23.8) | 1101 | (25.6) | 617 | (24.9) |
| University or graduate school | 13171 | (48.7) | 9958 | (49.1) | 1979 | (46.0) | 1234 | (49.8) |
| Occupation |  |  |  |  |  |  |  |  |
| Regular employees | 12575 | (46.5) | 9141 | (45.1) | 2220 | (51.7) | 1214 | (49.0) |
| Managers | 2541 | (9.4) | 1947 | (9.6) | 394 | (9.2) | 200 | (8.1) |
| Executives | 862 | (3.2) | 722 | (3.6) | 56 | (1.3) | 84 | (3.4) |
| Public service worker | 2810 | (10.4) | 2090 | (10.3) | 420 | (9.8) | 300 | (12.1) |
| Temporary workers | 2894 | (10.7) | 2160 | (10.7) | 489 | (11.4) | 245 | (9.9) |
| Freelances or professionals | 4454 | (16.5) | 3491 | (17.2) | 591 | (13.8) | 372 | (15.0) |
| Others | 900 | (3.3) | 710 | (3.5) | 128 | (3.0) | 62 | (2.5) |
| Company size |  |  |  |  |  |  |  |  |
| ≤9 employees | 6165 | (22.8) | 4865 | (24.0) | 830 | (19.3) | 470 | (19.0) |
| 10-49 employees | 4390 | (16.2) | 3243 | (16.0) | 755 | (17.6) | 392 | (15.8) |
| 50-99 employees | 2550 | (9.4) | 1879 | (9.3) | 437 | (10.2) | 234 | (9.4) |
| 100-499 employees | 5156 | (19.1) | 3822 | (18.9) | 893 | (20.8) | 441 | (17.8) |
| 500-999 employees | 1997 | (7.4) | 1433 | (7.1) | 355 | (8.3) | 209 | (8.4) |
| 1000-9999 employees | 4719 | (17.5) | 3472 | (17.1) | 741 | (17.2) | 506 | (20.4) |
| ≥10000 employees | 2059 | (7.6) | 1547 | (7.6) | 287 | (6.7) | 225 | (9.1) |
| Working hours per day |  |  |  |  |  |  |  |  |
| < 8h/d | 5334 | (19.7) | 4142 | (20.4) | 682 | (15.9) | 510 | (20.6) |
| 8≤ and <9h/d | 14848 | (54.9) | 11175 | (55.2) | 2252 | (52.4) | 1421 | (57.4) |
| 9 ≤ and <11h/d | 5541 | (20.5) | 4055 | (20.0) | 1045 | (24.3) | 441 | (17.8) |
| ≥11h/d | 1313 | (4.9) | 889 | (4.4) | 319 | (7.4) | 105 | (4.2) |
| Job contents questionnaire |  |  |  |  |  |  |  |  |
| Job demands | 30.1 | (5.9) | 29.5 | (5.7) | 32.5 | (6.1) | 30.4 | (5.6) |
| Job control | 63.4 | (11.5) | 63.9 | (11.6) | 60.9 | (11.6) | 63.9 | (10.9) |
| Supervisor support | 10.0 | (3.0) | 10.2 | (3.0) | 8.7 | (3.09) | 10.9 | (2.7) |
| Coworker support | 10.5 | (2.6) | 10.6 | (2.6) | 9.51 | (2.8) | 11.1 | (2.4) |
| Utrecht Work Engagement Scale-3 | 2.4 | (1.5) | 2.5 | (1.5) | 2.0 | (1.5) | 2.6 | (1.6) |

Control group is those who do not need the support for the work–treatment balance, unsupported group is those who need support but were not receiving it, and supported group is those who needed support and were actually receiving it.

### Comparison of the scores of the JCQ subscale among the groups

The scores of the JCQ subscale among the groups were compared according to the receipt state of the support for the work–treatment balance. In the supported group, the mean scores of the supervisor support and coworker support were the highest at 10.9 (2.7) and 11.1 (2.4) in the three groups. In the unsupported group, the mean score (SD) of the job demands was the highest of 32.5 (6.1), and those of the Job control, the supervisor support and the coworker support were the lowest of 60.9 (11.6), 8.7 (3.1), and 9.5 (2.8) (Table 1).

We statistically compared each subscale score of the JCQ among the groups by the receipt state of support for the work–treatment balance (Table 2). The job demand scores of the supported and unsupported groups were significantly higher than those of the control group in both age-sex and multivariate adjustment models (all p<0.001). There was no significant difference in the job control score between the supported and control groups in the sex-age-adjusted models; however, the job control score of the supported group was significantly higher than that of the control group in the multivariate model (p=0.013). The job control scores of the unsupported group were significantly lower than those of the control group in both models (both p<0.001).

**Table 2.** Comparison of the scores of the JCQ subscale and the UWES-3 among each group according to the receipt state of the support for the work–treatment balance.

| Parameters | Group | Sex-age adjusted |  |  | Multivariate *1 |  |  |
| --- | --- | --- | --- | --- | --- | --- | --- |
|  |  | Coefficients | 95%CI | p | Coefficients | 95%CI | p |
| JCQ subscale |  |  |  |  |  |  |  |
| Job demands | Supported | 0.81 | [0.57–1.05] | <0.001 | 0.78 | [0.55–1.01] | <0.001 |
|  | Unsupported | 2.92 | [2.73–3.11] | <0.001 | 2.69 | [2.51–2.88] | <0.001 |
|  | Control | reference |  |  | reference |  |  |
| Job control | Supported | 0.39 | [-0.09–0.86] | 0.111 | 0.57 | [0.12–1.02] | 0.013 |
|  | Unsupported | -2.78 | [-3.16– -2.41] | <0.001 | -2.32 | [-2.67– -1.96] | <0.001 |
|  | Control | reference |  |  | reference |  |  |
| Supervisor support | Supported | 0.70 | [0.58–0.82] | <0.001 | 0.66 | [0.54–0.78] | <0.001 |
|  | Unsupported | -1.45 | [-1.55– -1.36] | <0.001 | -1.42 | [-1.52– -1.32] | <0.001 |
|  | Control | reference |  |  | reference |  |  |
| Coworker support | Supported | 0.43 | [0.32–0.54] | <0.001 | 0.41 | [0.30–0.51] | <0.001 |
|  | Unsupported | -1.11 | [-1.20– -1.03] | <0.001 | -1.08 | [-1.16– -1.00] | <0.001 |
|  | Control | reference |  |  | reference |  |  |
| UWES-3 | Supported | 0.05 | [-0.01–0.11] | 0.125 | 0.06 | [-0.01–0.12] | 0.072 |
|  | Unsupported | -0.53 | [-0.57– -0.48] | <0.001 | -0.50 | [-0.55– -0.45] | <0.001 |
|  | Control | reference |  |  | reference |  |  |

CI: Confidence interval. JCQ, Job contents questionnaire, UWES-3: Utrecht Work Engagement Scale-3.
Control group is those who do not need the support for the work–treatment balance, unsupported group is those who need support but were not receiving it, and supported group is those who needed support and were actually receiving it.
\* The multivariate model was adjusted for age, sex, educational background, occupation, company size where participants work, working hours per day.

In the two models, the supervisor support scores of the supported group were significantly higher than those of the control group (both p<0.001), and those of the unsupported group were significantly lower than those of the control group (both p<0.001). There were no significant differences in the coworker support scores between the supported and control groups in the two models. The job control scores of the unsupported group were significantly lower than those of the control group in the two models (both p<0.001).

### Comparison of the scores of the UWES-3 among the groups

The scores of the UWES-3 among the groups were compared according to the receipt state of the support for the work–treatment balance. The mean UWES-3 score of the supported group was the highest at 2.6 (1.6), and that of the unsupported group was the lowest at 2.0 (1.5) in the three groups (Table 1). We statistically compared each UWES-3 score among the groups by the receipt state of support for the work–treatment balance (Table 2). There were no significant differences in the UWES-3 scores between the control and supported groups in the two models. The UWES-3 scores of the unsupported group were significantly lower than those of the control group in both models (both p<0.001).

## DISCUSSION

This study evaluated the relationship among the work–treatment balance, job stress, and work engagement. Regarding job demand–control, we found that the group of participants who received support for the work–treatment balance tended to be aware of lower job demand and higher job control than those who did not receive support. In particular, the group of participants who received support for work–treatment balance was aware of higher job control than those who did not need support for work–treatment balance when adjusting for multivariate. In a previous study of patients with inflammatory bowel disease, it was reported that work practices that negatively affect patients’ physical condition and health care behaviors and lack of consideration in the workplace are associated with decreased motivation to work and depression. ^17)^ Specific items of work considerations related to the work–treatment balance in Japan include the assignment of appropriate work practices, reduction of working hours such as limiting overtime work, change of work location, and consideration of hospital treatment and health care behaviors ^3)^. For those who receive support, these work considerations may reduce the psychological stress of work.

Regarding social support, we found that the group of participants who received support for work–treatment balance had a higher perception of supervisors and coworker support. For the work–treatment balance, the collaboration between the worker and the related parties (workplace staff, occupational physician, etc.) and the understanding of their coworkers toward the worker with the disease are important. ^3)^ We observed that those who received support for work–treatment balance belonged to large workplaces with more than 1,000 employees because these larger companies have better health management systems ^18)^. In particular, in Japan, workplaces with more than 1,000 employees employ dedicated occupational physicians, and their interventions and awareness-raising about the work–treatment balance may have a significant positive impact on increasing social support.

We found that the work engagement of the participants who have received support for the work–treatment balance is higher than that of those who need that support but are not receiving it, and is almost the same level as that of those who do not need that support. Based on the job demands-resources model (JD-R model), we believe that appropriate job demands and high social support as job resources contributed to the higher work engagement of the group. A meta-analysis examining the relationship between work engagement and outcomes reported that high work engagement has a positive impact on physical and mental health, organizational commitment, and job performance. ^19)^ We suggest that the approach to work–treatment balance could contribute significantly not only to the health condition of workers with diseases but also to their positive attitude toward work.

This study was conducted in December 2020 during the COVID-19 pandemic, and there are concerns that the psychological impact of the COVID-19 pandemic may worsen the health status of individuals. ^5, 6)^ We believe that work–treatment balance could be necessary during pandemics. For example, in Sweden, sick-leave rates almost doubled during March and April 2020, which was the first COVID-19 wave, compared with the previous year, suggesting that this increase in sick leave is largely due to prolonged COVID-19 symptoms ^20)^. COVID-19-related symptoms can be protracted and require intensive medical care ^21-23)^. In addition, it has been reported that the fear of COVID-19 infection or the government’s encouragement to avoid going out unnecessary may lead to refraining from taking action to seek medical care. ^9)^ This may lead to worsening of the disease and interruption of treatment. The COVID-19 epidemic has also caused major changes in the work system, such as the introduction of teleworking. ^24)^ The work–treatment balance will become increasingly important due to the major impact of the COVID-19 epidemic not only on human health but also on society.

This study has some limitations. First, because this study was an Internet-based survey, generalizability may be insufficient. We attempted to reduce bias in recruiting participants. Second, this study is a cross-sectional study, and the causal relationship between work–treatment balance and job stress or work engagement is not clear. Third, the concrete disease diagnoses of the participating workers receiving the support of the work–treatment balance is unknown. In Japan, the proportion of workers with mental health disorders is higher than that of brain and heart diseases ^3)^, and it cannot be denied that there was a bias in these results. Fourth, this study was conducted during a COVID-19 epidemic, and we cannot deny the possibility that this may have modified these results. Further research should be conducted at normal times.

## CONCLUSION

This study has shown an association among support for the work–treatment balance, job stress, and work engagement in Japan. Those who received support for the work–treatment balance showed lower job stress and higher work engagement. Therefore, providing work–treatment balance for workers could have a positive impact on their mental health.

## Data Availability

Due to the nature of this research, the participants of this study did not consent to their data being publicly shared; hence, supporting data will not be made available.

## Acknowledgments

The current members of the CORoNaWork Project, in alphabetical order, are as follows: Dr. Yoshihisa Fujino (present chairperson of the study group), Dr. Akira Ogami, Dr. Arisa Harada, Dr. Ayako Hino, Dr. Hajime Ando, Dr. Hisashi Eguchi, Dr. Kazunori Ikegami, Dr. Kei Tokutsu, Dr. Keiji Muramatsu, Dr. Koji Mori, Dr. Kosuke Mafune, Dr. Kyoko Kitagawa, Dr. Masako Nagata, Dr. Mayumi Tsuji, Ms. Ning Liu, Dr. Rie Tanaka, Dr. Ryutaro Matsugaki, Dr. Seiichiro Tateishi, Dr. Shinya Matsuda, Dr. Tomohiro Ishimaru, and Dr. Tomohisa Nagata. All members were affiliated with the University of Occupational and Environmental Health, Japan.

## Author contributions

KI created the questionnaire, analyzed the data, and wrote the manuscript. HA and AO reviewed the manuscript, analyzed the data, and provided advice on interpretation. HE,□MT,□ST,□TN,□and SM reviewed the manuscript. YF reviewed the manuscript and contributed to the overall survey planning, questionnaire creation, and securing funding for research.

## Ethical approval

This study was approved by the ethics committee of the University of Occupational and Environmental Health, Japan (reference No. R2-079).

